# Child Suicide Rates During the COVID-19 pandemic in England

**DOI:** 10.1101/2021.07.13.21260366

**Authors:** David Odd, Tom Williams, Louis Appleby, David Gunnell, Karen Luyt

## Abstract

**Background:** There is concern about the impact of COVID-19, and the control measures to prevent the spread, on children’s mental health. The aim of this work was to identify if there had been a rise of childhood suicide during the COVID pandemic; using data from England’s National Child Mortality Database (NCMD).

**Method:** Child suicide rates between April to December 2020 were compared with those in 2019 using negative binomial regression models, and characteristics compared. In a subset (1^st^ January to 17^th^ May 2020) further characteristics and possible contributing factors were obtained.

**Results:** A total of 193 likely childhood deaths by suicide were reported. There was no evidence overall suicide deaths were higher in 2020 than 2019 (RR 1.09 (0.80-1.48), p=0.584) but weak evidence that the rate in the first lockdown period (April to May 2020) was higher than the corresponding period in 2019 (RR 1.56 (0.86-2.81), p=0.144). Characteristics of individuals were similar between periods. Restriction to education and other activities, disruption to care and support services, tensions at home and isolation appeared to be contributing factors.

**Limitations:** As child suicides are fortunately rare, the analysis is based on small numbers of deaths with limited statistical power to detect anything but major increases in incidence.

**Conclusion:** We found no consistent evidence that child suicide deaths increased during the COVID-19 pandemic although there was a concerning signal they may have increased during the first UK lockdown. A similar peak was not seen during the following months, or the second lockdown.

## Introduction

The COVID-19 pandemic and the public health measures to control its spread have had a major impact on the everyday lives of people around the world. In England alone, by the 8^th^ of March 2021, over 4,223,232 people had tested positive for COVID-19, and over 124,566 COVID related deaths had been identified [1]. There is concern that the physical distancing measures taken to control the spread of COVID-19 may have had an adverse impact on the mental health and wellbeing of children and young people and, as a result, about the possibility of a rise in child suicide [2–4]. While suicide rates in children are low compared to other demographic groups, the rate in the under 20s has been rising in England and Wales since around 2010 [5]. Young people in their late teens also have the highest rate of non-fatal self-harm, a key suicide risk factor, and this appears to have risen in recent years [6]. Multiple factors contribute to an individual’s risk of suicide [7], while additional stressors during the pandemic may include fears that a family member or oneself will develop COVID-19, the impact of bereavement, isolation, loneliness and loss of social supports, disruptions to care and support and fears about accessing it, school closure and exam disruption, and exposure to domestic violence and family tensions [3,8]. In the UK the government instigated a ‘lockdown’ process on the 23rd of March, similar to the ‘stay-at-home’ orders enacted in other countries, with a raft of recommendations, including closing schools and businesses, in order to reduce the spread of COVID-19. This was lifted on the 1st of June, although a variable amount of social restrictions remained until the 6th of November when a second stay-at-home order was enacted. Early analysis of suicide surveillance data from England’s National Child Mortality Database raised a signal of concern about child suicide rates in the first 8 weeks of the pandemic [9]. More recently, an analysis of Japanese suicide data for children and young adults (<20 years) identified a rise in suicides in the second, but not the first period of school closure in Japan [10].

### Aims

The aim of this work was to identify if there had been a rise of childhood suicide during the COVID pandemic and the subsequent social lockdown, and identify any pandemic-related factors or changes in the characteristics of children dying by suicide.

## Method

The NCMD collects data from the 58 Child Death Overview Panels (CDOPs) that review the deaths of all children who die before their 18^th^ birthday in England [11]. There is a legal responsibility for CDOPs to notify NCMD of any death of someone aged under 18 years within 48 hours of it occurring, using an electronic system. The NCMD commenced data collection on 1^st^ April 2019. NCMD notification data includes details on deaths referred to the coroner; such deaths do not appear in official statistics until after an inquest has occurred, often many months later.

The National Child Mortality Database (NCMD) responded to the pandemic by accelerating its notification and analysis service to develop a real-time surveillance system. The notification details provided for all deaths reported to the NCMD, up to the 31^st^ of December 2020 were reviewed and categorised by 4 people (3 paediatricians and one NCMD Manager with CDOP expertise) to identify likely suicide deaths. Where there was not full agreement, the deaths were reviewed by each member of the team again. Deaths where there was still disagreement were then reviewed by a researcher with expertise in suicide research (DG), blind to the date of death. In keeping with the approach used in previous research [12], this final review categorised the likelihood that these deaths were by suicide as: high, moderate, low or unclear, based on all the available information. In addition to the information provided in the notification details field, additional data from the notification form were reviewed:

- Sex of individual (female, male, other (including not known))
- Ethnic Group (Asian or Asian British, Black or Black British, Mixed, Other, Unknown, White)
- Age at death
- Deprivation decile of the child’s home address using the Index of Multiple Deprivation (IMD) [13]. Decile of deprivation is calculated using 7 main domains (income, employment, education, health, crime, access to housing and services, and living environment) and is calculated from the child’s postcode to a granularity of around 1500 people.
- Free text description of the method of suicide and (of varying detail) the circumstances surrounding the death (including information on apparent precipitants of the likely suicide, social environment, and history of contacts with services).

To assess the rate of suicide during the pandemic, and to take account of any possible seasonal differences in trends and risk factors [14], deaths occurring in three pandemic periods of 2020 (Period 1: First National Lockdown: 1^st^ April 2020 to 31^st^ May 2020; Period 2: eased restrictions between lockdowns: 1^st^ June 2020 to 5^th^ November 2020; Period 3: Second National Lockdown in 2020 6^th^ November to 31^st^ December 2020) were compared with the same periods in 2019. Due to absence of data prior to April 2019, the first week of the national lockdown (23^rd^ March to 31^st^ March 2020) was excluded from this analysis. In a second analysis the number of deaths in 2020 was described for the four phases of the pandemic in England; Pre-Lockdown (1^st^ January to 22^nd^ March), the 1^st^ National Lockdown (23^rd^ March to 31^st^ May), during the subsequent period of less severe social distancing measures (1^st^ June to 5^th^ November) and then the 2^nd^ National Lockdown (6^th^ November to 31^st^ December).

To investigate if the characteristics of the children dying of suicide changed during the pandemic we initially compared the available characteristics of them. Next, in a subset of deaths occurring in 2020 (1^st^ of January to the 17^th^ of May), a bespoke questionnaire was sent to each CDOP that had notified a likely suicide death to gather further information. Both the pre- and post-lockdown questionnaires requested information about any history the individuals had of past or current contact with mental health or social services and related psychiatric diagnoses. In addition, the post-lockdown questionnaire also requested information on whether there was any evidence that lockdown and/or school closure contributed to the child’s death and whether any difficulties had been identified in accessing mental health or social services during lockdown. Investigation of the deaths was at an early stage, so information provided in the questionnaires was preliminary and only partially complete. For all the post-lockdown deaths enough information was provided to enable analysis, but in two pre-lockdown deaths we had insufficient information available and therefore these deaths were excluded from the qualitative analysis.

### Statistical Analysis

To assess the rate of suicide during the pandemic our primary analysis was based on those deaths between the 1^st^ of April 2019 and the 31^st^ of December 2020 (n=193) where it was considered suicide was highly or moderately likely to be the cause of death. Data was collapsed to provide frequency counts of events per day with events during 2020 compared with the same period of 2019. A negative binomial regression model with an estimated underlying at-risk population of 12,023,568 (based on Office for National Statistics mid-2019 estimates for children under the age of 19 years in England)[15] was used to compare rate of suicide in each of the three periods of the pandemic with the equivalent periods in 2019. The models were then repeated for suicides that occurred throughout 2020, comparing the three periods outlined above (extending the first period of lockdown to include 23^rd^ March to 31^st^ March) with suicide rates in the pre-lockdown period of 2020 (1^st^ January to 22^nd^ March). We then carried out a sensitivity analysis, repeating the analyses including the deaths where there was initial disagreement on cause, and the subsequent expert opinion was that the chance of suicide was unclear or low (n=14).

To investigate if the characteristics of the children dying of suicide changed during the pandemic the characteristics of deaths in the two periods were compared using Fishers exact test for categorical data and Mann-Whitney U for age and deprivation decile. We did not have complete data for all deaths, and in addition for this analysis, as numbers were small, sex coded as ‘Other’ was not included and ethnicity was coded as “white” or “other”.

All analysis was performed using Stata Version 16. Data were analysed on the 08/03/2021. Data is presented as Rate Ratio (RR) (95% confidence interval (CI)), number (%) or median (interquartile range (IQR)) as appropriate. P-values lower than 0.05 were considered evidence of statistical significance.

## Results

### Number of Suicides

Between 1^st^ April 2019 and 31^st^ December 2020 (641 days) a total of 193 likely childhood deaths by suicide were reported to NCMD. Suicide was the cause of 3% (n=193) of all child deaths reported over this period; but accounted for 9% (n=45) of all deaths between 10 and 14 years and 25% (n=146) of deaths of 15-17 year olds. Using ONS data for population size,[15] this approximates to 0.8 suicides per 100,000 children per year for those 10 to 14 years old, and 4.5 suicides per 100,000 children per year for those 15 to 17 years old. Due to the small number of suicides, the overall rate per day varied across the time period investigated, although the highest rate was seen during the first lockdown (Figure 1).

**Figure 1.**
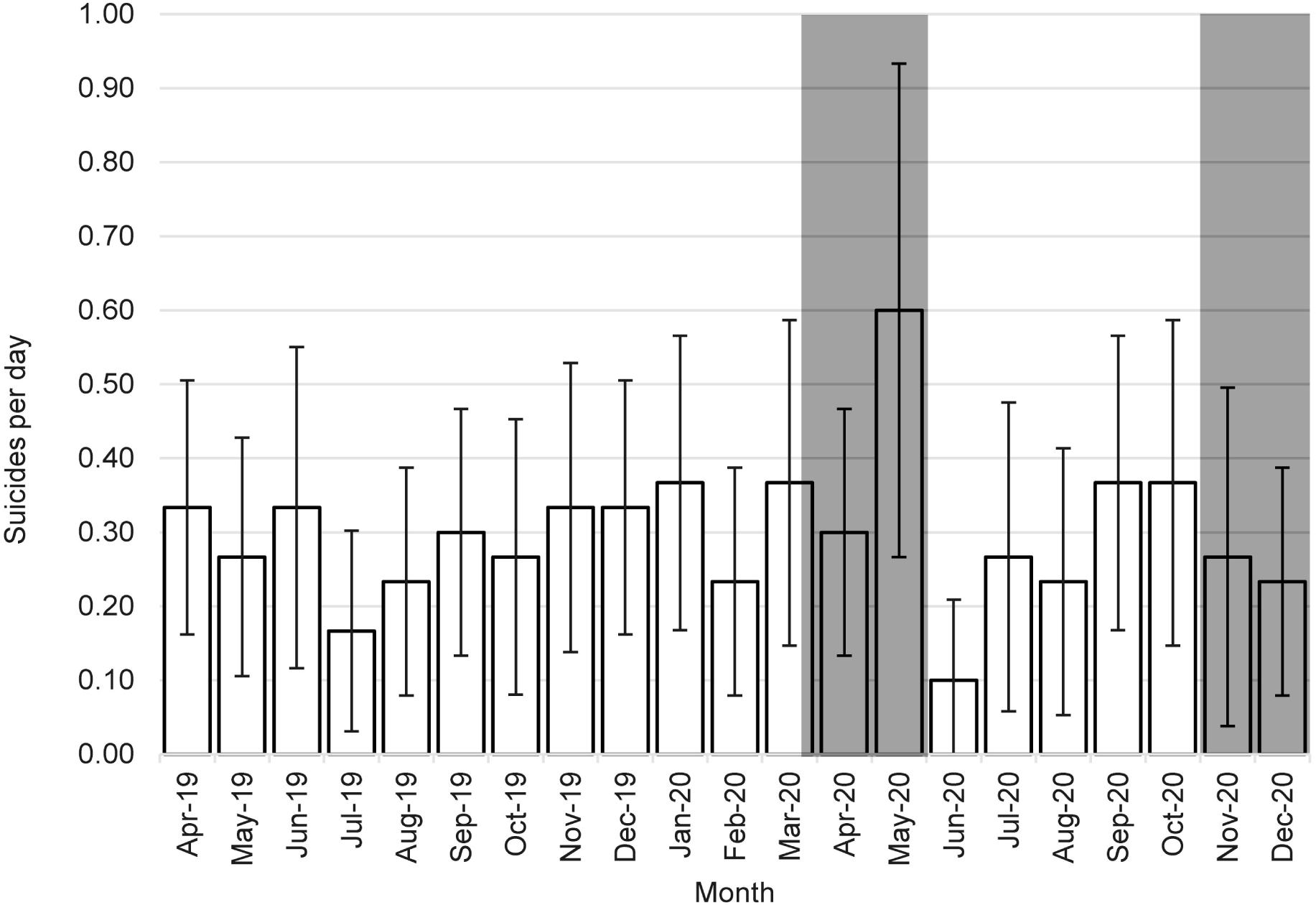
Mean suicides deaths per day over each calendar month from April 2019 to December 2020. Shaded areas represent periods of National Lockdown.

A total of 163 likely suicide deaths were identified during the periods of 1^st^ April to 31^st^ December across both 2020 and 2019 (a total of 550 days) and overall there was little evidence that suicide deaths were higher in 2020 than 2019 (85 deaths in 2020 vs. 78 in 2019; RR 1.09 (0.80-1.48), p=0.584) (Table 1). While not reaching conventional levels of statistical significant, there was weak evidence that the rate in the first lockdown (Period 1) was higher than the corresponding period the previous year (RR 1.56 (0.86-2.81), p=0.144); although no association was seen in the immediate post-lockdown period (Period 2; RR 1.10 (0.72-1.69), p=0.663) or in the second lockdown (Period 3; RR 0.65 (0.32-1.31), p=0.227). Findings were similar when suicide rates in the 3 pandemic periods (March 23-31 December 2020) were compared with rates in the weeks leading up to the pandemic (January 1^st^ 2020-March 22^nd^ 2020). A final model repeated the analysis to also include deaths where there was initial disagreement on cause, and the subsequent expert opinion was that the chance of suicide was unclear or low (Additional 14 deaths). This produced similar results to the main analysis (2020 vs 2019: RR 1.11 (0.82-1.49)).

**Table 1.** Relative Rate of Child Suicide between time periods of interest.

| **2020 vs 2019 (April-Dec)** | **Dates** | **Days** | **Number of Deaths** |  | **Relative Rate (95% CI)** | **p-value** |
| --- | --- | --- | --- | --- | --- | --- |
|  |  |  | **2019** | **2020** |  |  |
| Deaths in 2020 vs 2019 | 1st April to 31st December | 275 | 78 | 85 | 1.09 (0.80-1.48) | 0.584 |
| Split by time periods |  |  |  |  |  |  |
| During First Lockdown | 1<sup>st</sup> April to 31<sup>st</sup> May | 61 | 18 | 28 | 1.56 (0.86-2.81) | 0.144 |
| Between Lockdowns | 1<sup>st</sup> June to 5<sup>th</sup> November | 158 | 40 | 44 | 1.10 (0.72-1.69) | 0.663 |
| Second Lockdown | 6<sup>th</sup> November to 31<sup>st</sup> December | 56 | 20 | 13 | 0.65 (0.32-1.31) | 0.227 |
| **Deaths only in 2020** | **Dates** | **Days** | **Number of Deaths** |  | **Relative Rate (95% CI)** | **p-value** |
| Before lockdown | 1<sup>st</sup> Jan to 22<sup>nd</sup> March | 82 | 26 |  | Ref | - |
| During First Lockdown | 23<sup>rd</sup> March 2020 to 31<sup>st</sup> May | 70 | 32 |  | 1.44 (0.85-2.44) | 0.173 |
| Between Lockdowns | 1<sup>st</sup> June to 5<sup>th</sup> November | 158 | 44 |  | 0.88 (0.54-1.44) | 0.604 |
| During Second Lockdown | 6<sup>th</sup> November to 31<sup>st</sup> December | 56 | 13 |  | 0.73 (0.37-1.44) | 0.364 |

Results are Relative Rate (95% CI)

### Characteristics of Deaths

There was no evidence that the sex, ethnicity, or IMD decile of the likely suicide deaths differed in the 3 phases of social restrictions in 2020 compared the same period of 2019 (Table 2). There was weak evidence of a lower age for suicide deaths in 2020 than 2019 (16.2 (14.5-17.2) vs 16.8 (14.9-17.6) years, p=0.077) and while an initial review of the deaths indicated a possible higher proportion of children aged under 15 years[9] the proportion of deaths younger than 15 years was similar between 2020 and 2019 (23 (27.1%) vs 20 (25.6%), p=0.861).

**Table 2.**
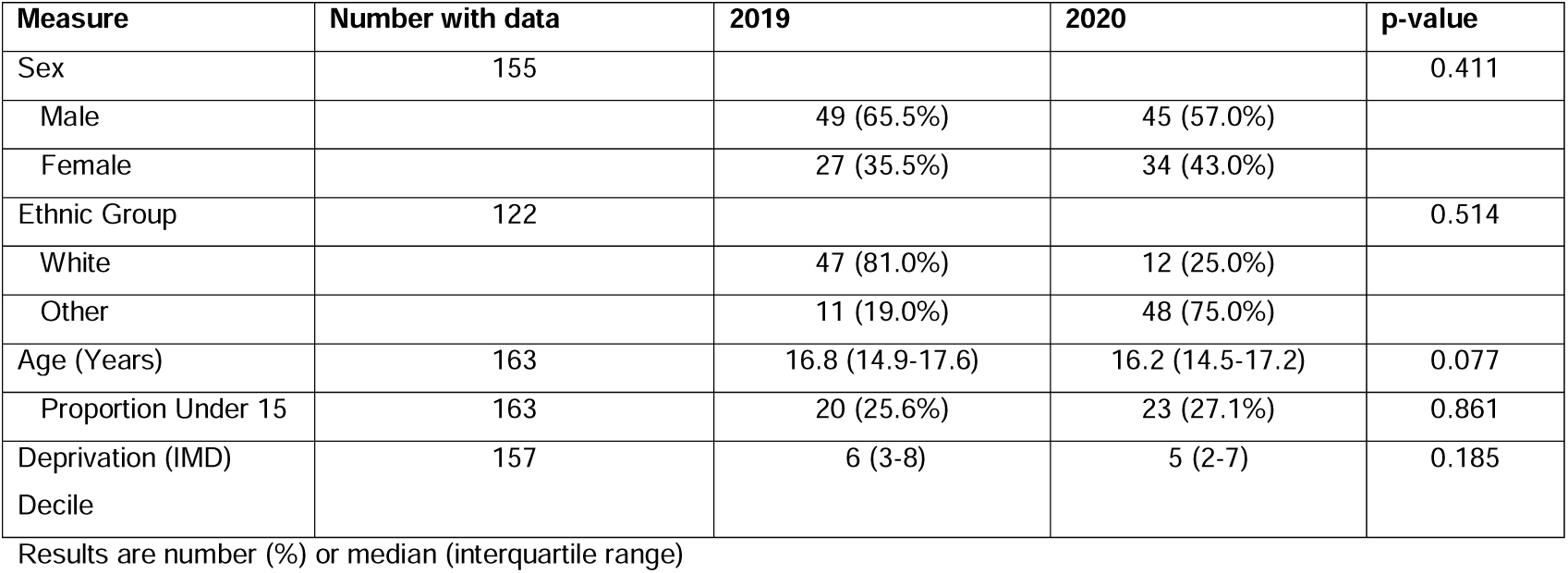
Characteristics of child suicides from 1 April to 31^st^ of December 2020 vs. the same period in 2019.

### Review of subset of deaths occurring between 1 January 2020 and 17th May 2020

In both time periods the most common method used for likely suicide was hanging (17 out of 25 before, and 20 out of 25 after lockdown). We were unable to obtain detailed information on two deaths before lockdown and so the review of these was limited to 24 in the pre lockdown period (1^st^ January to 22^nd^ March 2020) and 25 in the during lockdown period (23^rd^ March to 17^th^ of May 2020).

Pre lockdown deaths: Of the 24 deaths reviewed, in eight (33%) the individuals were specified as being under current follow-up with mental health services or social care and a further 6 (25%), children had previous contact with mental health services or social care. Altogether, 14 (58%) were reported as having some current or past contact with services. In 12 this contact was with mental health services; 6 (25% overall) of these children had a diagnosis of ASD, ADHD or both.

Post lockdown deaths: Of the 25 deaths, nine (36%) were specified as being under current follow up with mental health services or social care and a further eight (32%) children had previous contact with mental health services or social care. Altogether, 17 (68%) were reported as having some current or past contact with services. In 13 this contact was with mental health services; 6 (24% overall) of these children had a diagnosis of ASD, ADHD or both. COVID-19 related factors were reported by CDOPs to have contributed to the death in 12 (48%) of the 25 deaths identified during lockdown; contributors included: restrictions to education and other activities, disruption to care and support services, tensions at home and isolation.

## Discussion

We found no strong or consistent evidence that child suicide deaths increased during the COVID-19 pandemic although there was a concerning signal that child suicide deaths may have increased in the first period of lockdown in 2020, this was not seen after lockdown restrictions were eased, and did not appear to reoccur in the second lockdown. Nevertheless there are concerns that the mental health of children has been affected by the pandemic[4] and a continued focus on this age group is needed.

Amongst the likely suicide deaths reported during lockdown, restrictions to education and other activities, disruption to care and support services, tensions at home and isolation appeared to be important factors; although reporting may reflect increased vigilance during lockdown. In keeping with this finding, a recent study of adolescents admitted to a psychiatric unit in the US during the pandemic, patients with COVID-related suicidal thoughts or behaviours reported higher levels of stressors related to missing special events, financial problems, conflict at home and changes in living circumstances [8]. A UK national study of suicides amongst 10-19 year olds in 2014-2016 suggested that academic pressures were an important factor in 32% of suicides, bereavements in 25% and social isolation or withdrawal in 21%[16], and all such factors are likely to contribute to risk during the pandemic.

A recent analysis of the impact of the pandemic on suicide rates (all ages) in 21 countries up to July 2020 found no overall rise[17]; and indeed a number of areas appeared to have experienced falls in suicide deaths. Nevertheless there are particular concerns about marginalised groups and children, and a recent analysis of data from Japan indicated that whilst suicides amongst those aged under 20 years did not increase during the first period of national restrictions and school closure, there was evidence of rise during the second wave of restrictions and school closure [10].

Data from other studies on the incidence of non-fatal self-harm amongst children during the pandemic are similar. There were falls in primary care presentation for self-harm in 10-17 year olds in the early months of the pandemic although rates returned to pre-pandemic levels by September 2020 [18]. However, community surveys have suggested a rise in mental ill-health [4].

The COVID-19 pandemic comes at a time when there is growing concern over rising suicide and self-harm rates in young people in the UK [5,19,20]. While rates are low in children compared to other demographic groups, childhood rates in the UK have been rising since around 2010 [5], and similar trends have been seen in young adulthood suicide in other countries [21,22].

The causal processes contributing to each suicide death are complex. Suicide is often associated with multiple factors including adversity in early childhood, bullying, personal and parental mental illness, exposure to suicidal behaviour in others and genetic vulnerability[7]/ During this period of lockdown, known factors such as isolation [23], loss of social support, disruptions to care and support and potential exam disruption [3], or direct anxiety regarding viral illness, may become greater; as may limitations in accessing social, mental health, and other services while the NHS and other providers try to adapt to new ways of working. However, it is possible that for some people social distancing may lead to an improvement in their symptoms (e.g. those with school phobias). However, while other reports have raised concerns about those with mental health needs during this pandemic[24], the proportion of children in contact with services was similar amongst children who died in the pre and post lockdown periods (33% vs 36%).

Particular concerns have been raised about the impact of school closures and physical distancing measure on children with ASD or ADHD [25]. We found no evidence that their risk increased during the pandemic, although children with these diagnoses comprised around a quarter of all the likely suicide deaths in our detailed nested study of suicides pre vs. post lockdown. Our work adds to existing concerns regarding self-harm and suicide in this group [26].

### Limitations

There are several limitations to our analysis. As child suicides are fortunately rare, the analysis is based on small numbers of deaths, meaning we had limited statistical power to detect anything but major increases in incidence. In addition, categorisation of each death was based on limited data, and most deaths in this analysis are awaiting full CDOP review. It is possible that some deaths may be unreported, due to the CDOPs themselves not being notified that the death has occurred, or reported too late to be included in this work; although all deaths in 2020 so far were reported within 2 weeks of the date of death.

## Supporting information

Ethics and Consent Statement

## Data Availability

The data that support the findings of this study are available on request from the corresponding author [KL] given appropriate approvals. The data are not publicly available due to containing information that could compromise the privacy.

## Funding

The National Child Mortality Database (NCMD) Programme is commissioned by the Healthcare Quality Improvement Partnership (HQIP) as part of the National Clinical Audit and Patient Outcomes Programme (NCAPOP). HQIP is led by a consortium of the Academy of Medical Royal Colleges, the Royal College of Nursing, and National Voices. Its aim is to promote quality improvement in patient outcomes. HQIP holds the contract to commission, manage and develop the National Clinical Audit and Patient Outcomes Programme (NCAPOP), comprising around 40 projects covering care provided to people with a wide range of medical, surgical and mental health conditions. NCAPOP is funded by NHS England, the Welsh Government and, with some individual projects, other devolved administrations and crown dependencies www.hqip.org.uk/national-programmes. DG is supported by the NIHR Biomedical Research Centre at University Hospitals Bristol and Weston NHS Foundation Trust and the University of Bristol, England. The views expressed in this publication are those of the author(s) and not necessarily those of the NHS, the National Institute for Health Research or the Department of Health and Social Care.

## Ethics and Consent Statement

The NCMD legal basis to collect confidential and personal level data under the Common Law Duty of Confidentiality has been established through the Children Act 2004 Sections M - N, Working Together to Safeguard Children 2018 (https://consult.education.gov.uk/child-protection-safeguarding-and-family-law/working-together-to-safeguard-children-revisions-t/supporting_documents/Working%20Together%20to%20Safeguard%20Children.pdf) and associated Child Death Review Statutory & Operational Guidance https://assets.publishing.service.gov.uk/government/uploads/system/uploads/attachment_data/file/859302/child-death-review-statutory-and-operational-guidance-england.pdf). The NCMD legal basis to collect personal data under the General Data Protection Regulation (GDPR) without consent is defined by GDPR Article 6 (e) Public task and 9 (h) Health or social care (with a basis in law).

## Acknowledgements

Supported by: The NCMD Team (Vicky Sleap, Kate Hayter, Sylvia Stoianova, Lacia Ashman, Peter Fleming)

Statistical Advice from Professor Chris Metcalfe (University of Bristol), Dr Matt Spittal (University of Melbourne) and Dr Duleeka Knipe (University of Bristol)

We thank all Child Death Overview Panels (CDOPs) who submitted data for the purposes of this report and all child death review professionals for submitting data and providing additional information when asked.

## Authors’ contributions

David Odd: I declare that I participated in the study concept and design, analysis and interpretation of data, drafting and review of the manuscript and that I have seen and approved the final version.

Tom Williams: I declare that I participated in the study design, contributed to interpretation of data, drafting the manuscript; and that I have seen and approved the final version.

Louis Appleby: I declare that I participated in the study design, contributed to analysis and interpretation of data, drafting the manuscript; and that I have seen and approved the final version.

David Gunnell: I declare that I participated in the study design, contributed to analysis and interpretation of data, drafting the manuscript; and that I have seen and approved the final version.

Karen Luyt: I declare that I participated in the study design, contributed to analysis and interpretation of data, drafting the manuscript; and that I have seen and approved the final version.

## Declarations of interest

David Odd, Karen Luyt and Tom Williams have no conflicts of interest. Louis Appleby chairs the Department of Health’s National Suicide Prevention Strategic Advisory Group (England). David Gunnell is a member of the Department of Health’s National Suicide Prevention Strategic Advisory Group (England), and is a member of Samaritans Policy, Partnerships and Research Committee and Movember’s Global Advisory Committee.

## Notes

### Author Declarations

The NCMD legal basis to collect confidential and personal level data under the Common Law Duty of Confidentiality has been established through the Children Act 2004 Sections M - N, Working Together to Safeguard Children 2018 (https://consult.education.gov.uk/child-protection-safeguarding-and-family-law/working-together-to-safeguard-children-revisions-t/supporting_documents/Working%20Together%20to%20Safeguard%20Children.pdf) and associated Child Death Review Statutory & Operational Guidance https://assets.publishing.service.gov.uk/government/uploads/system/uploads/attachment_data/file/859302/child-death-review-statutory-and-operational-guidance-england.pdf). The NCMD legal basis to collect personal data under the General Data Protection Regulation (GDPR) without consent is defined by GDPR Article 6 (e) Public task and 9 (h) Health or social care (with a basis in law). In addition we have obtained confirmation of exemption to require additional ethical approval from the Chair of Central Bristol NHS REC (uploaded).

### Summary of Updates

In error, we omitted to thank a academic who gave advice. We have added her name to the acknowledgements. No other changes.

