## Supplementary material for "Child Suicide Rates During the COVID-19 pandemic in England": Ethics and Consent Statement

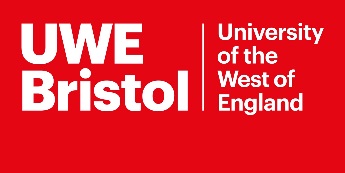

Faculty of Health & Applied
Sciences
Glenside Campus
Blackberry Hill
Stapleton
Bristol	BS16 1DD

Dear Publisher

This is just to explain that the ethical permission status of the study below:

**National Child Mortality Database: Real-time child mortality surveillance during the COVID-19 pandemic**

The database is set up as a part of statutory duty to record child mortality.

The NCMD has a legal basis to collect confidential and personal level data under the Common Law Duty of Confidentiality has been established through the Children Act 2004 Sections M - N, Working Together to Safeguard Children 2018 ([https://consult.education.gov.uk/child-protection-safeguarding-and-family-law/working-together-to-safeguard-children-revisions-t/supporting_documents/Working_Together_to_Safeguard_Children.pdf](https://consult.education.gov.uk/child-protection-safeguarding-and-family-law/working-together-to-safeguard-children-revisions-t/supporting_documents/Working%20Together%20to%20Safeguard%20Children.pdf))
and associated Child Death Review Statutory & Operational Guidance 
<https://assets.publishing.service.gov.uk/government/uploads/system/uploads/attachment_data/file/859302/child-death-review-statutory-and-operational-guidance-england.pdf>). The NCMD legal basis to collect personal data under the General Data Protection Regulation (GDPR) without consent is defined by GDPR Article 6 (e) Public task and 9 (h) Health or social care (with a basis in law).

This database is therefore set up so that researchers can access the data that is routinely collected. External researchers might apply to access this data but in this instance the team are carrying out the research that this data base was set up to facilitate.

This study therefore does not require NHS permissions or NHS ethical approval.

Hope that is helpful but please contact me if you require further clarification.

Yours sincerely

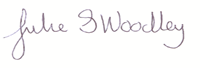

Dr Julie Woodley PhD,MSc,DCR(R),TDCR(R),HDCR(R),FETC,ARRT (R).

Chair Central Bristol NHS REC

Rm 2K01 Glenside Campus

Faculty of Health and Social Wellbeing

Blackberry Hill

Stapleton

Bristol BS16 1AD (+44 (0)117 3288528)
